# A pilot study to investigate the safety and feasibility of antiretroviral therapy for Alzheimer’s disease (ART-AD)

**DOI:** 10.1101/2024.02.26.24303316

**Authors:** A. Campbell Sullivan, Gabrielle Zuniga, Paulino Ramirez, Roman Fernandez, Chen-Pin Wang, Ji Li, Lisa Davila, Kristine Pelton, Sandra Gomez, Claira Sohn, Elias Gonzalez, Marisa Lopez-Cruzan, David A. Gonzalez, Alicia Parker, Eduardo Zilli, Gabriel A. de Erausquin, Sudha Seshadri, Sara Espinoza, Nicolas Musi, Bess Frost

## Abstract

Retrotransposons are viral-like DNA sequences that constitute approximately 41% of the human genome. Studies in *Drosophila,* mice, cultured cells, and human brain indicate that retrotransposons are activated in settings of tauopathy, including Alzheimer’s disease, and causally drive neurodegeneration. The anti-retroviral medication 3TC (lamivudine), a nucleoside analog reverse transcriptase inhibitor, limits retrotransposon activation and suppresses neurodegeneration in tau transgenic *Drosophila,* two mouse models of tauopathy, and in brain assembloids derived from patients with sporadic Alzheimer’s disease. We performed a 24-week phase 2a open-label clinical trial of 300 mg daily oral 3TC (NCT04552795) in 12 participants aged 52-83 years with a diagnosis of mild cognitive impairment due to suspected Alzheimer’s disease. Primary outcomes included feasibility, blood brain barrier penetration, effects of 3TC on reverse transcriptase activity in the periphery, and safety. Secondary outcomes included changes in cognition and fluid-based biomarkers of neurodegeneration and neuroinflammation. All participants completed the six-month trial; one event of gastrointestinal bleeding due to a peptic ulcer was reported. 3TC was detected in blood and cerebrospinal fluid (CSF) of all participants, suggestive of adherence to study drug and effective brain penetration. Cognitive measures remained stable throughout the study. Glial fibrillary acidic protein (GFAP) (*P*=0.03) and Flt1 (*P*=0.05) were significantly reduced in CSF over the treatment period; Aβ42/40 (*P*=0.009) and IL-15 (*P*=0.006) were significantly elevated in plasma. While this is an open label study of small sample size, the significant decrease of some neurodegeneration- and neuroinflammation-related biomarkers in CSF, significantly elevated levels of plasma Aβ42/40, and a trending decrease of CSF NfL after six months of 3TC exposure suggest a beneficial effect on subjects with mild cognitive impairment due to suspected Alzheimer’s disease. Feasibility, safety, tolerability, and central nervous system (CNS) penetration assessments further support clinical evaluation of 3TC in a larger placebo-controlled, multi-dose clinical trial.

## INTRODUCTION

While the estimated global prevalence of Alzheimer’s disease dementia is frequently cited as greater than 50 million people^1^ and costs over a trillion dollars in the United States annually, these estimates fail to capture those in the early stages of the disease process. When looking across the Alzheimer’s disease continuum, from those in the ‘preclinical’ stages to those with advanced dementia, the global estimates balloon to approximately 416 million people^2^, accounting for roughly 22% of individuals over the age of 50 worldwide.

Given the high prevalence rates and mounting economic and societal burdens, efforts to develop disease-modifying treatments for Alzheimer’s disease have accelerated. The FDA recently approved two treatments targeting the removal of abnormal aggregates of Aβ in the brain. While promising, these anti-amyloid treatments have yielded limited clinical results in patients with Alzheimer’s disease^3^. Similar lackluster findings have been reported for efforts targeting tau removal in the brain^4,5^. While strategies to clear aggregated Aβ and tau remain promising, these proteins begin to accumulate in the brain decades prior to a typical clinical diagnosis of Alzheimer’s disease^6^. The nodes within the sequela of toxic events induced by Aβ or tau deposition thus provide novel pharmacological targets for mitigating disease progression.

We^7–9^ and others^10–12^ have reported that pathogenic forms of tau drive neurotoxicity through the activation of retrotransposons in *Drosophila,* cell, and mouse models of tau pathology. Retrotransposons are “selfish” genetic elements that compose over 40% of the human genome. Such elements are thought to have derived from viral infections that occurred over the course of evolution; intact retrotransposons are similar to retroviruses in that they have the ability to replicate their DNA via an RNA intermediate and insert the new copy into the genome^13^. In addition to generating insertional mutations, retrotransposons can activate the innate immune system through the production of viral-like double stranded RNAs (dsRNA), extrachromosomal circular DNAs (eccDNA), and proteins^14–19^, and can cause DNA double strand breaks as a consequence of failed insertion^20^. Transcriptional analyses of post-mortem human brain identify retrotransposon-encoded transcripts, particularly of the “endogenous retrovirus” class, that are elevated in Alzheimer’s disease and progressive supranuclear palsy, a “primary” tauopathy^7^, and suggest that such activation is a consequence of pathogenic forms of tau^10^. In line with these findings, longitudinal analysis of blood isolated from patients with Alzheimer’s disease further reveals a significant increase in retrotransposon transcripts prior to phenoconversion from normal cognition to cognitive impairment^21^. Work in model organisms of tauopathy further demonstrate that retrotransposon activation is a causal factor driving neurodegeneration and neuroinflammation^7,11,12,22,23^.

The reverse transcriptase inhibitor 3TC effectively reduces retrotransposon activation and neurotoxicity in *Drosophila*^7^, mouse^11,12^, cell^11^, and brain assembloid^24^ models of tauopathy and Alzheimer’s disease. 3TC is an anti-retroviral medication approved by the FDA in the 1990’s to treat HIV/AIDS and chronic hepatitis B and continues to be prescribed as a mono- or combination therapy for retroviral infection. 3TC is considered safe, is widely prescribed, and has few clinically-relevant pharmacological interactions due to its low metabolic clearance, minimal binding to plasma protein, and no detectable effects on liver function^25–28^. In addition to the positive effects of 3TC in laboratory models of Alzheimer’s disease and related tauopathies, analysis of health insurance databases in the United States indicates that use of nucleoside reverse transcriptase inhibitors such as 3TC is associated with lower incidence of Alzheimer’s disease^29^. Given this compelling preclinical data and the encouraging safety profile of 3TC, we conducted the first clinical trial of 3TC in older adults with predementia Alzheimer’s disease.

## RESULTS

### Study design, participant characteristics and drug adherence

Primary outcomes of this 24-week open-label phase 2a trial were to evaluate feasibility, CNS penetration, effects on circulating levels of reverse transcriptase activity, and safety and tolerability of the nucleoside analog reverse transcriptase inhibitor 3TC in subjects with mild cognitive impairment due to suspected Alzheimer’s disease. Secondary outcomes explored cognitive performance and changes in CSF and plasma biomarkers related to neurodegeneration and neuroinflammation from baseline to post-treatment. 22 candidates were pre-screened to identify 12 participants aged 50-99 years with a Clinical Dementia Rating (CDR) of 0.5 and Mini-Mental State Examination (MMSE) score between 24 and 30. Eligibility criteria are included in **Extended Data Table 1**. Participants were enrolled between March 18, 2021 and September 15, 2022. Baseline participant characteristics are summarized in **Table 1**.

**Table 1.** Study participant characteristics.

| CHARACTERISTIC | N |
| --- | --- |
| Number of participants | 12 |
| Average age, years (min, max) | 69.4 (52, 83) |
| Age group (years) |  |
| ≤ 65 | 3 (25%) |
| 65 - 74 | 5 (41.7%) |
| 75 - 84 | 4 (33.3%) |
| Sex |  |
| Female | 9 (75%) |
| Male | 3 (25%) |
| Race |  |
| White, non-Hispanic | 9 (75%) |
| White, Hispanic | 1 (8.3%) |
| African American | 1 (8.3%) |
| Asian | 1 (8.3%) |
| Education, years (SD) | 15.3 (2.4) |
| Past medical history |  |
| ≤1 Comorbidities | 2 (16.7%) |
| 2+ Comorbidities | 10 (83.3%) |
| Dyslipidemia | 9 (75%) |
| Prediabetes | 4 (33.3%) |
| Diabetes | 2 (16.7%) |
| Hypertension | 6 (50%) |
| Hypothyroidism | 2 (16.7%) |
| Duration of symptoms prior to trial |  |
| ≤1 year | 1 (8.3%) |
| 1-2 years | 2 (16.7%) |
| 3+ years | 2 (16.7%) |
| Unknown | 7 (58.3%) |
| Average BMI, kg/m<sup>2</sup> (SD) | 25.2 (4.6) |
| Concurrent treatment with donepezil | 7 (58.3%) |

Following the initial screening visit, participants were subject to a comprehensive neuropsychological exam (**Extended Data Table 2**), blood draw (visit 1) and lumbar puncture (visit 2) prior to initiation of 300 mg daily oral 3TC (visit 3) (**Table 2**). Participants returned to the clinic at weeks 8, 16, and 24 of treatment (visits 4-6) to complete medication checks, physical examinations, and blood draw. At week 24 of treatment, participants completed a post-treatment comprehensive neuropsychological exam, blood draw (visit 6), and lumbar puncture (visit 7). One month after the final dose of medication, participants returned to the clinic for a final safety assessment and disenrollment (visit 8). All study participants completed the trial. CSF was not collected from three of 12 participants due to procedure failure, procedure-related pain, or patient-reported illness.

**Table 2.** Study visit schedule.

|  | Screening | Visit 1 | Visit 2 | Visit 3 | Visit 4 | Visit 5 | Visit 6 | Visit 7 | Visit 8 |
| --- | --- | --- | --- | --- | --- | --- | --- | --- | --- |
|  | Safety measurements |  |  |  |  |  |  |  |  |
| Vitals/labs | X | X |  | X | X | X | X |  | X |
| Height/weight | X | X |  | X |  |  | X |  |  |
| Adverse event review |  |  |  | X | X | X | X |  | X |
|  | Neuropsychological testing |  |  |  |  |  |  |  |  |
| CDR, MMSE | X |  |  |  |  |  |  |  |  |
| Neuropsychological battery |  | X |  |  |  |  | X |  |  |
|  | Clinical procedures |  |  |  |  |  |  |  |  |
| Blood draw | X | X |  |  | X | X | X |  |  |
| Lumbar puncture |  |  | X |  |  |  |  | X |  |
|  | Study medication |  |  |  |  |  |  |  |  |
| Administer in clinic |  |  |  | X | X | X |  |  |  |
| Compliance/tolerability |  |  |  |  | X | X | X |  |  |

3TC is absorbed rapidly after oral administration, reaching peak serum concentrations within 1.5-3 hours^30^. Previous analysis of 3TC brain penetration reports that the concentration of 3TC in CSF is 4-8% of serum concentrations 2-4 hours following treatment. Blood draws and lumbar punctures for 3TC analyses were performed within 24 hours of drug administration. 3TC was detected in plasma and CSF in all participants at all time points (**Extended Data Fig. 1A, B**), indicative of brain penetration and in line with full medication adherence as reported by all participants and caregivers at each visit. Plasma and CSF 3TC levels varied widely among participants and study visits, likely due to the short half-life of 3TC.

Some retrotransposons encode a viral-like reverse transcriptase that acts to convert retrotransposon RNA into new DNA copies. We quantified reverse transcriptase activity in plasma collected from participants at baseline and after 24 weeks of 3TC treatment using a modified version of the EnzCheck Reverse Transcriptase Assay, a fluorescent dye-based assay that quantifies the ability of a given sample to generate RNA:DNA heteroduplexes from a poly(A) template^31^. Levels of reverse transcriptase activity in plasma at baseline and following 24 weeks of 3TC treatment were variable among participants and did not significantly differ in response to treatment (**Extended Data Fig. 2**).

### Safety and treatment-emergent adverse events

3TC is widely used in antiretroviral therapy regimens due to its potent and long-lasting antiviral efficacy and low toxicity. Adverse events defined as unfavorable medical occurrences related to participation in the research were reviewed at visits 3-6 and at the follow-up visit one month following the last administration of 3TC. There was one event of gastrointestinal bleeding due to a peptic ulcer in a participant who was taking daily aspirin. Two participants (ART-AD-11 and ART-AD-22) reported a COVID-19 infection during the treatment period. The remaining participants reported no treatment-emergent adverse events. Serial laboratory studies did not reveal signs of liver toxicity (e.g. elevated liver enzymes, bilirubin, hypoglycemia, anemia) or cytopenia, which are previously-reported 3TC-related adverse events^32–34^. Given that weight loss has been reported in previous studies of 3TC-containing antiretroviral regimens^35^, we compared standing weights at baseline and after 24 weeks of treatment. Subjects lost 1.83 kg on average, ranging from 8.62 kg loss (ART-AD-02) to 3.99 kg gain (ART-AD-22), after daily 3TC for 24 weeks. No deaths occurred within the treatment period or one month after treatment.

### Effects of 3TC on cognition and functional status

Following 24 weeks of treatment, we measured cognitive change from baseline using a comprehensive neuropsychological battery (**Table 3**). There was no significant change in cognition among participants from baseline to follow-up assessment. In addition to exploring post-treatment changes in cognition across the test battery, secondary aims assessed changes in the Preclinical Alzheimer Cognitive Composite (PACC-5) score. Although typically relegated to individuals with prodromal and asymptomatic disease, the PACC-5 was included given its sensitivity to Alzheimer’s disease-specific cognitive change^36^. Similarly, no statistically significant changes were observed. While we detect a trend for small decreases in MMSE (P=0.06) and PACC-5 (P=0.07) with treatment, this trend resolves after participants with a COVID-19 infection during the treatment period are removed from the analysis (MMSE, P=0.2; PACC-5, P=0.2). MMSE and PACC-5 scores for each participant are included in **Extended Data Fig. 3**.

**Table 3.** Cognitive assessments. IQR=interquartile range. *P* value is based on Wilcoxon signed rank test with continuity correction. N=12.

| DOMAIN | TEST | BASELINE<br>MEDIAN (IQR) | POST-TREATMENT<br>MEDIAN (IQR) | P-value |
| --- | --- | --- | --- | --- |
| Global | **MMSE** | 27 (25, 29) | 25 (23, 28) | 0.06 |
| AD Composite | **PACC-5 (Z-score)** | -1.59 (-2, -0.59) | -1.59 (-2, -1.2) | 0.07 |
| Attention &<br>Working Memory | **Digits Forward** | 6 (5, 7) | 6 (5, 7) | 0.8 |
|  | **Digits Backward** | 4 (4, 5) | 4 (4, 5) | 0.2 |
| Verbal List Learning | **HVLT Learning** | 15 (14, 18) | 16 (13, 21) | >0.9 |
|  | **HVLT Recall** | 0 (0, 4) | 0 (0, 4) | >0.9 |
|  | **HVLT Disc** | 6 (2, 9) | 6 (3, 9) | >0.9 |
| Visual Learning | **BVMT Learning** | 9 (5, 11) | 8.5 (6, 10) | 0.8 |
|  | **BVMT Recall** | 2 (0, 3) | 2 (1, 3) | 0.8 |
|  | **BVMT Disc** | 4 (3, 5) | 4 (3, 5) | 0.4 |
| Story Memory | **Logical Memory I** | 22 (18, 25) | 18 (16, 24) | 0.3 |
|  | **Logical Memory II** | 6 (2, 9) | 4 (0, 8) | 0.8 |
|  | **Logical Memory Rec** | 16 (14, 18) | 16 (14, 16) | 0.3 |
| Episodic Memory | **FCSRT Delay Free** | 2 (0, 6) | 2 (0, 6) | 0.7 |
|  | **FCSRT Delay Cued** | 9 (6, 10) | 10 (8, 11) | 0.8 |
| Naming | **BNT** | 29 (28, 29) | 29 (27, 30) | 0.4 |
| Verbal Fluency | **Animal Naming** | 14 (13, 17) | 15 (13, 17) | 0.4 |
|  | **Supermarket** | 20 (16, 26) | 19 (16, 22) | 0.11 |
|  | **S Words** | 11 (10, 14) | 11 (10, 13) | 0.7 |
|  | **P Words** | 12 (9, 13) | 12 (10, 14) | 0.08 |
| Executive<br>Functioning | **Trails A Time** | 44 (38, 64) | 49 (32, 56) | 0.7 |
|  | **Trails B Time** | 114 (94, 205) | 105 (92, 280) | 0.4 |
|  | **Digit-Symbol Coding** | 46 (34, 49) | 39 (37, 47) | >0.9 |

### Effects of 3TC on neurodegeneration-related biomarkers

We analyzed the effects of 24-week 3TC treatment on biomarkers that have been previously linked to neurodegeneration^37^. Commercial SIMOA (Single Molecule Array) assays were performed on an HD-X platform (Quanterix) to quantify NfL, GFAP, Aβ42, Aβ40, and pTau181 in CSF (**Fig. 1A-D**) and plasma (**Fig. 1E-F**). Despite our small sample size, we find that GFAP is significantly reduced in CSF after 24 weeks of 3TC treatment, suggestive of reduced astrocyte reactivity and neuroinflammation due to 3TC. We also detect a significant increase in Aβ42/40 in plasma after 24 weeks of 3TC treatment, suggestive of decreased amyloid pathology in the brain due to 3TC.

**Figure 1.**
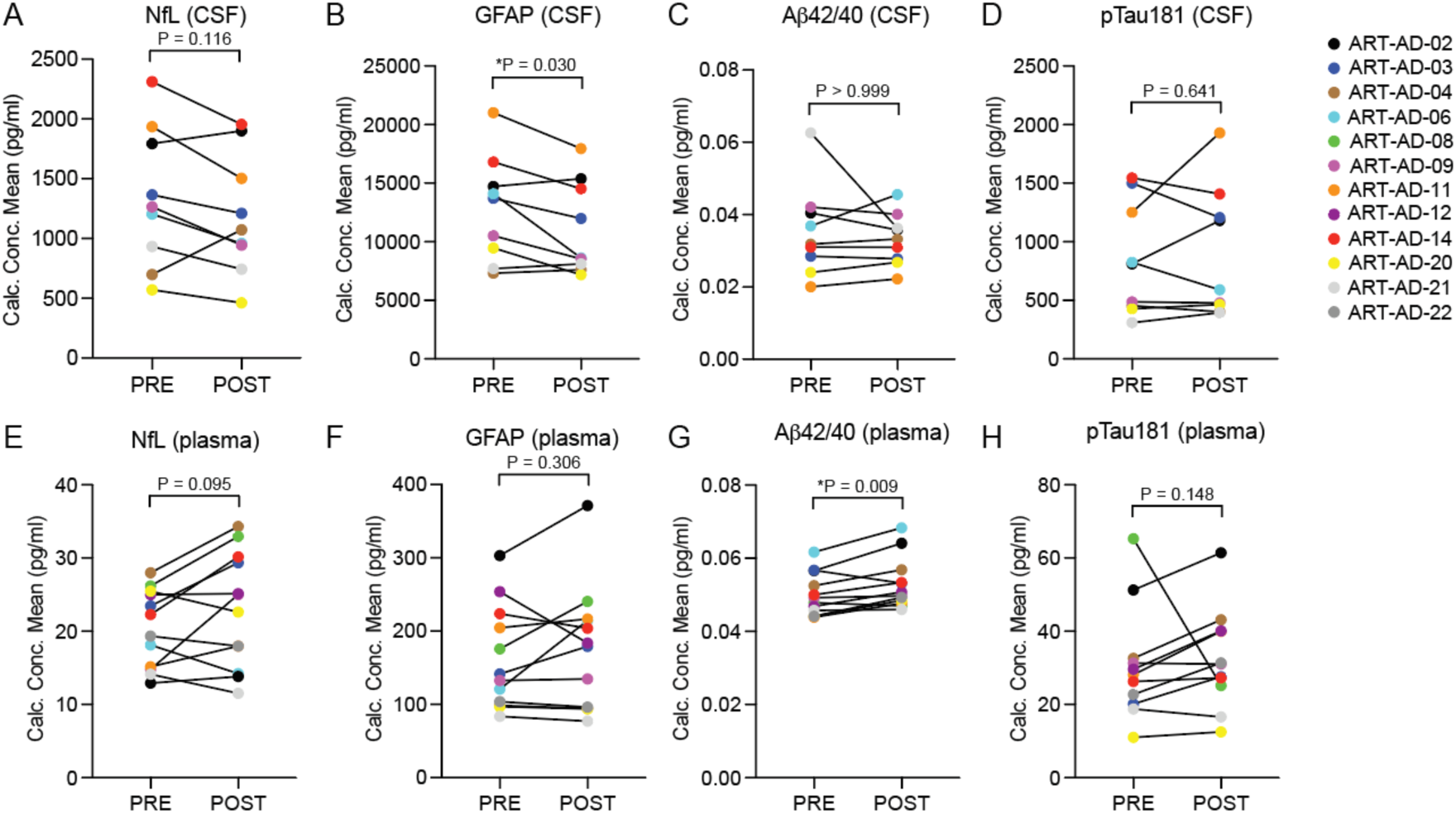
Quantification of biomarkers associated with neurodegeneration in CSF and plasma. Quanterix was used to detect the indicated proteins in CSF (A-D) and plasma (E-H) of participants at baseline and after 24 weeks of 3TC treatment. CSF, N=9; Plasma, N=12. Normality testing was based on the POST-PRE difference for each target; *P* values are based on two-sided paired sample t-test (normal distribution) or Wilcoxon matched-pairs signed rank test (non-normal distribution).

### Effects of 3TC on biomarkers of neuroinflammation

The Mesoscale Discoveries (MSD) V-PLEX Neuroinflammation Panel 1 was used to analyze CSF and plasma levels of 37 biomarkers that are associated with a neuroinflammatory response. Biomarkers that were either not detected in more than three participants or those in which the % coefficient of variation (%CV) exceeded 15 were removed from the analysis. This left 21 high-confidence targets for analysis in CSF and 30 high-confidence targets for analysis in plasma (**Table 4, Extended Data Figs. 4, 5**). In CSF, we detect a reduction in Flt1 from baseline to post-treatment (*P*=0.05). In plasma, we detect an increase in IL-15 from baseline to post-treatment (*P*=0.006). The remaining neuroinflammatory biomarkers were not significantly changed over the course of treatment.

**Table 4.**
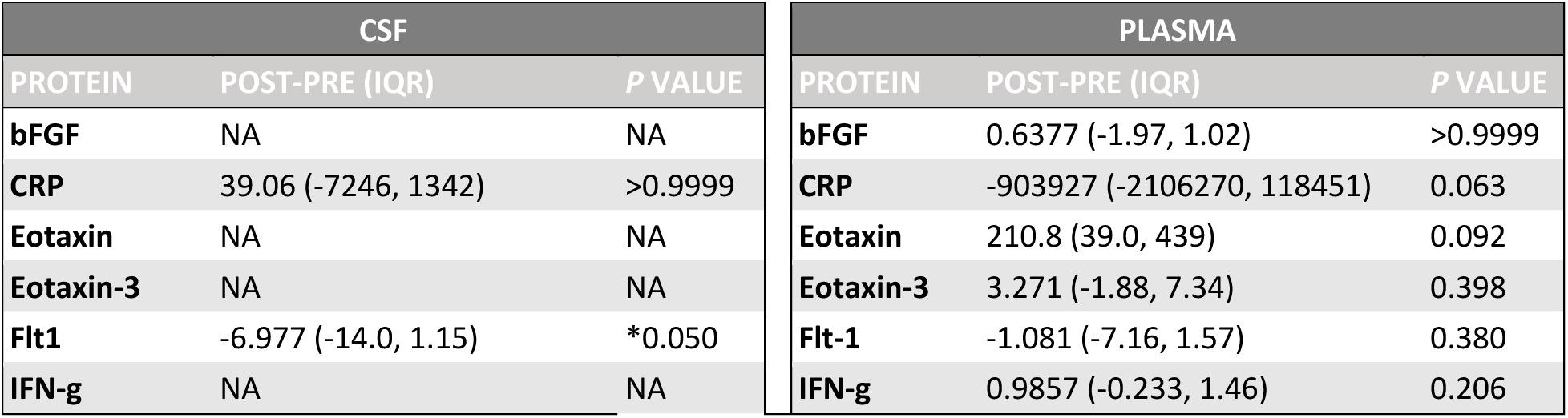
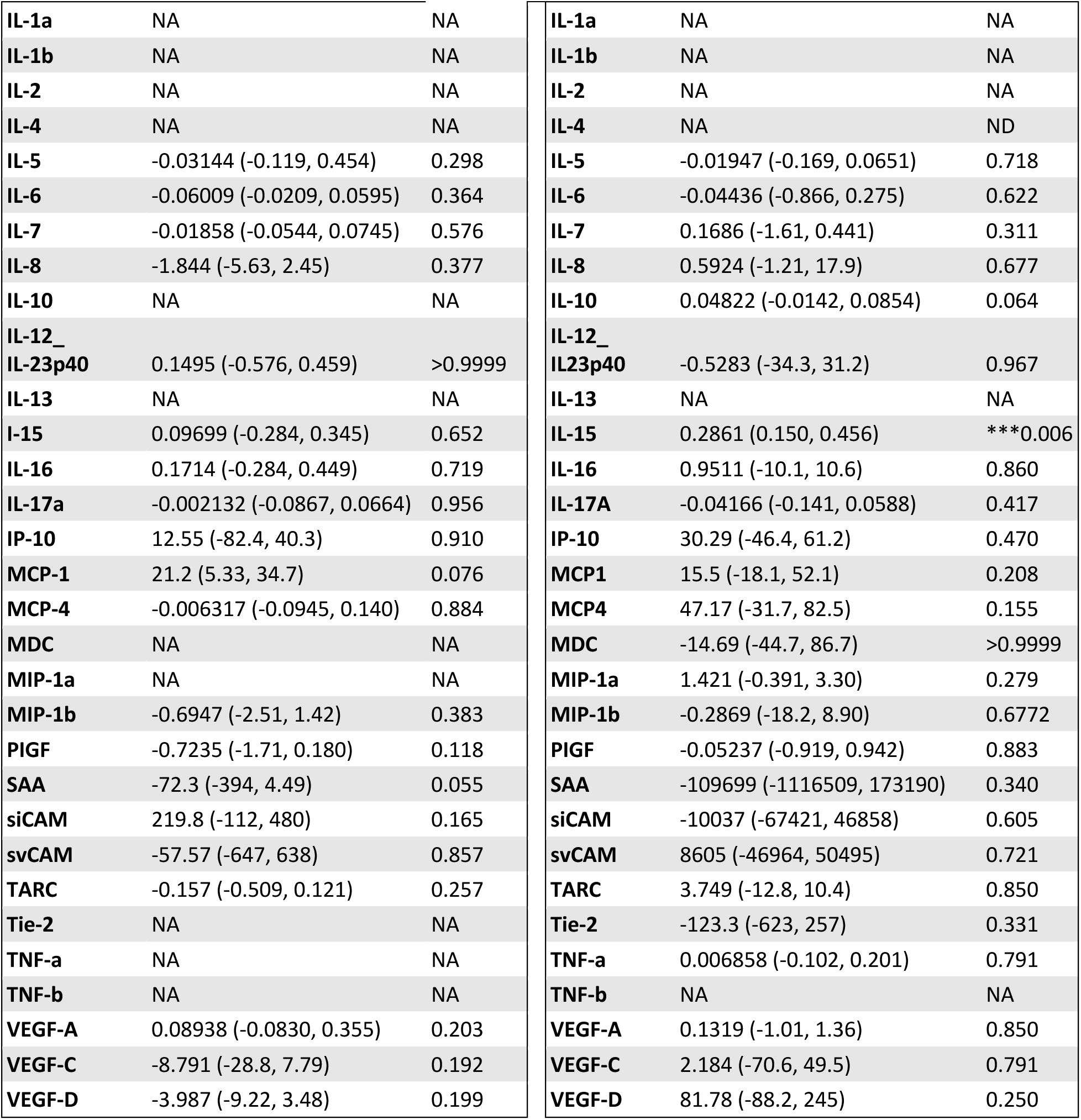
CSF and plasma-based inflammatory biomarker analysis. A value of “NA” is included for targets in which three or more participants had undetectable values or %CV values exceeding 15. POST-PRE values are in pg/ml. CSF, N=9; Plasma, N=12. Normality testing was based on the POST-PRE difference for each target; *P* values are based on two-sided paired sample t-test (normal distribution) or Wilcoxon matched-pairs signed rank test (non-normal distribution). IQR=Interquartile Range.

## METHODS

### Study design and intervention

ART-AD is an investigator-initiated, single center, open-label phase 2a clinical trial. Enrolled subjects were stable on current medications for at least eight weeks prior to initiating 300 mg oral 3TC (McKesson Medical Supplies, product #1121405) daily at the initial visit. This dose was maintained throughout the 24-week study with excellent medication adherence as assessed by pill count. All 12 subjects received the same dose and frequency. Subjects presented to the clinic at regular visits according to the protocol outlined in **Table 2**. This study was conducted according to US and international standards of Good Clinical Practice (FDA Title 21 part 312). The study protocol was approved by the University of Texas Health San Antonio Institutional Review Board and was registered at ClinicalTrials.gov NCT04552795. Participant identification numbers were not known to any individuals outside of the research group.

### Fluid collection and processing

Fasting peripheral venous blood was collected into BD Paxgene® DNA tubes, BD EDTA coated tubes, and BD serum separator tubes (SST) for DNA, hematology, and chemistry analyses, respectively. Peripheral blood mononuclear cells (PBMC) were isolated from plasma using BD CPT tubes that contain Ficoll-Hypaque. Plasma and PBMCs were aliquoted into polypropylene tubes and stored at −80 °C. Complete blood count with differential and platelets, coagulation, metabolic, and lipid panels, and hemoglobin A1c were analyzed by LabCorp.

Lumbar punctures were performed at the Glenn Biggs Institute for Alzheimer’s and Neurodegenerative Diseases after overnight fasting. 2 ml of CSF was collected into a 10 ml polypropylene tube via gravity drip using a spinal needle. Resulting CSF was centrifuged to remove cellular debris, then aliquoted into polypropylene tubes and stored at −80 °C.

### Neuropsychological assessment

All participants completed comprehensive neuropsychological assessment (see **Extended Data Table 2**). These evaluations were completed at baseline (prior to the initiation of 3TC), and then again following 24-weeks of continued treatment. The PACC-5 score was calculated as a mean normative z-score across five measures, including MMSE, Logical Memory Delayed Recall, Digit-Symbol Coding Test, Category Fluency, and Free and Cued Selective Reminding Test.

### HPLC/MS/MS chromatography

3TC ((−)-L-2ʹ,3ʹ-dideoxy-3ʹ-thiacytidine) and internal standard (3-guanidinopropionic acid (3GPA)) were obtained from Millipore Sigma. All other HPLC-grade reagents were purchased from Thermo Fisher Scientific. All solutions were prepared with Milli-Q water. A 3TC super stock was prepared in methanol at a concentration of 1 mg/ml and stored in aliquots at −80 °C. The working stock solution of 3TC was prepared each day from the super stock at a concentration of 100 μg/ml to spike the calibrators.

Reference calibrators were prepared by spiking control plasma to concentrations of 0, 5, 10, 25, 50, 100, 500, 1,000 ng/ml of 3TC, and control CSF to concentrations of 0, 2, 10, 50, 100, 500, 1,000, 5,000 ng/ml of 3TC. An identical concentration of internal standard, 10 µl (100 µg/ml 3-GPA), was spiked into each of the calibrators and blinded samples. 100 µl of mobile phase B (0.1% formic acid in acetonitrile) was added to each tube, vortexed thoroughly, and spun for 5 minutes at 17,000xg. 10 µl of the supernatant was injected into the HPLC with MS detection.

The LC/MS/MS system consisted of a Shimadzu SIL 20A HT autosampler, two LC-20AD pumps, and an AB Sciex API 4000 tandem mass spectrometer with turbo ion spray. The LC analytical column was an ACE Excel C18-PFP (75 x 3.0 mm, 3 micron) purchased from Mac-Mod Analytical and was maintained at 24 °C during the chromatographic runs using a Shimadzu CT-20A column oven. Mobile phase A containing 0.1% formic acid dissolved in water was run at 0.255 ml/min. Mobile phase B consisted of 0.1% formic acid dissolved in 100% HPLC grade acetonitrile and run at 0.045 ml/min. The total flow rate of the mobile phase was 0.3 ml/min. 3TC and the internal standard, 3-GPA, were eluted isocratically with a 5-minute run. 3TC eluted at 3.69 min, while 3-GPA eluted at 3.25 min. Drug transitions were detected in positive mode at m/z 230 à 112 for 3TC and m/z 132 à 72 for 3-GPA. The ratio of 3TC peak area to the internal standard peak area for each blinded sample was compared against a linear regression of the ratios obtained by the calibration peak areas to quantify 3TC.

### Reverse transcriptase activity

Quantification of reverse transcriptase activity was performed using the EnzCheck reverse transcriptase assay kit (Invitrogen, E22064). Frozen plasma samples were thawed on ice and centrifuged at 2,000 rpm for 10 minutes to remove debris. Protein within the resulting supernatant was concentrated using 50K molecular weight cut off concentrators (Thermo Fisher Scientific, 88539). Reverse transcriptase reactions were run with diluted samples in freshly prepared enzyme dilution buffer (50 mM Tris-HCl, 20% glycerol, 2 mM DTT, pH 8) to microplate wells with and without the reaction mixture containing template/primer solution in polymerization buffer. The reaction was incubated for six hours at room temperature, after which 200 mM EDTA was added to stop the reaction. The RNA-DNA heteroduplexes formed in the reaction were detected via PicoGreen; fluorescence was measured using a microplate reader (Promega GloMax Discover). To correct for nonspecific binding, the fluorescence level of the sample (plasma sample with reaction mixture) was subtracted from the background (plasma sample without reaction mixture). The corrected data was used to calculate reverse transcriptase activity based on the reverse transcriptase standard curve.

### Fluid biomarker analysis

Plasma and CSF samples were analyzed in the Fluid Biomarkers Laboratory at the Brown University Center for Alzheimer’s Disease Research. Samples and controls were run in duplicate and calibrators were run in triplicate. The measurements were performed blinded to diagnosis and clinical data in one round of experiments using one batch of reagents. Biomarkers related to neurodegeneration and neuroinflammation were quantified from the same CSF or plasma sample for each subject. The mean inter-assay and intra-assay CV values were below 10% for all markers. Samples with CV values greater than 15% were removed from the analysis. Targets lacking data for more than three participants (due to lack of detection or CV > 15%) were not included in the analysis.

#### Measurement of Aβ40, Aβ42, NfL, GFAP and pTau181

Plasma and CSF levels of the analytes of interest were determined by commercial SIMOA (Single Molecule Array) assays on an HD-X platform (Quanterix). Levels of Aβ42, Aβ40, GFAP, and NfL were quantified using the Neurology 4-plex E kit (103670; Quanterix) in CSF (400-fold dilution) and plasma (4-fold dilution). Levels of pTau181 were quantified using the pTau181 Advantage kit version 2.1 (104111; Quanterix) in CSF (10-fold dilution) and plasma (4-fold dilution).

Samples were thawed briefly on wet ice. Immediately after thawing, CSF samples used for Aβ40, Aβ42, NfL and GFAP assays were diluted 400x in N4PE CSF Sample Diluent. All calibrators, controls, and samples were then incubated at room temperature for one hour. Samples and internal controls were centrifuged at 10,000 x g for five minutes. All plasma samples were diluted 4x onboard using the provided sample diluent. CSF samples were diluted at the bench and run neat. A four-parameter logistic curve fit, 1/y2 weighted was used for GFAP, NfL and pTau181 while a 5-paramter logistic curve fit, 1/y2 weighted was used for Aβ40 and Aβ42. Two control samples of known concentration (high-control and low-control) provided in each kit as well as two internal controls of pooled CSF and plasma were included as quality control.

#### Measurement of inflammatory markers

Inflammatory proteins were measured according to manufacturer’s instructions using the V-PLEX Neuroinflammation Panel 1 (K15210D; Meso Scale Diagnostics) on the Meso QuickPlex SQ 120 (MSD). Plasma and CSF samples were diluted 2-fold for the V-plex Pro-inflammatory panel 1 (IFN-γ, IL-1β, IL-2, IL-4, IL-6, IL-8, IL-10, IL-13 and TNF-α), Cytokine Panel 1 (1L-1α IL-5, IL-7, IL-12/23p40, IL-15, IL-16, IL-17A, TNF-β and VEGFA) and Angiogenesis panel 1 (VEGF-C, VEGF-D, Tie-2, Flt-1, PIGF and bFGF). Plasma and CSF samples were diluted 4-fold for the Chemokine Panel 1 (Eotaxin, MIP-1β, Eotaxin-3, TARC, IP-10, MIP-1α, MCP-1, MDC and MCP-4). CSF samples were diluted 5-fold and plasma samples were diluted 1000-fold for the Vascular Injury Panel 1 (SAA, CRP, VCAM-1 and ICAM-1). Levels of MCP-4 (CSF), MIP-1a (plasma), TARC (CSF), IL-17a (CSF), IL-10 (plasma), IL-5 (plasma), IL-17a (plasma) fell below the lower limit of detection (LLOD) during the initial screen. The assay was repeated for those markers only using a 2-fold dilution with an overnight incubation at 4°C^38^.

Samples were thawed briefly on wet ice and centrifuged at 2,000 x g for three minutes. After washing of the precoated plates with Phosphate Buffered Saline plus 0.05% Tween-20, 50 μl of calibrator control and diluted sample was added to each well. Plates were sealed and incubated for two hours at room temperature or overnight at 4 °C with shaking at 700 rpm. After washing, 25 μl of detection antibody solution was added to each well and the plate was incubated for two hours at room temperature with shaking at 700 rpm for all assays except Vascular Injury Panel 1, which was incubated for one hour. The plates were washed again followed by the addition of 150 μl Read Buffer T to each well. Plates were read immediately with the exception of Chemokine Panel 1, which was incubated for ten minutes at room temperature prior to reading. A four-parameter logistic curve fit, 1/y2 weighted was used for all assays. Control samples of known concentration were included as quality control.

### Statistical analyses

#### Cognitive assessments

Given the small sample size and skewed distribution, non-parametric (Wilcoxon matched-pairs signed rank test) tests were used to examine cognitive change following 24-weeks of 3TC. SPSS was used for statistical analyses of cognitive data.

#### Fluid biomarker analyses

For each target, the range of POST-PRE values among all participants was tested for normality using D’Agostino’s K-squared test. A paired two-sided t-test was used to calculate *P* values for targets in which POST-PRE values passed the normality test (alpha = 0.05), while *P* values for targets that failed the normality test were calculated using the Wilcoxon matched-pairs signed rank test. A *P* value of less than or equal to 0.05 was considered significant. Multiple comparison testing was not performed due to the exploratory nature of the study and to avoid false negatives. GraphPad Prism was used for statistical analyses and figure preparation.

## Discussion

Activation of retrotransposons has been reported in post-mortem human tissue and fluids from patients with Alzheimer’s disease and related tauopathies^7,10,21^, as well as in *Drosophila* ^7,9,22^, mouse^8,11,12^, and cell culture-based models^11,24^. Mechanistically, data suggest that retrotransposon activation is a consequence of the effects of pathogenic forms of tau on chromatin architecture and small RNA-mediated silencing of retrotransposon transcripts^7^. Studies in *Drosophila* ^7^ and mouse models^11,12^ of tauopathy indicate that tau-induced retrotransposon activation causally drives neurodegeneration and is pharmacologically targetable using the nucleoside analog reverse transcriptase inhibitor 3TC. In the current study, we provide results of the first clinical trial testing the effects of 3TC in participants with mild cognitive impairment due to suspected Alzheimer’s disease.

We find that 24-week treatment with 300 mg daily oral 3TC (the standard dosing regimen for HIV and hepatitis B) is safe and well-tolerated, and that 3TC effectively penetrates the blood brain barrier in this population. 3TC is absorbed rapidly after oral administration, with an absolute bioavailability of 82% in adults^39^. 3TC was detected in plasma and CSF of all participants, indicating adherence to the study drug. 3TC levels were variable among participants and between study visits. The variability of 3TC detected in plasma and CSF among participants likely reflects the short half-life of 3TC. In future studies, we suggest controlled timing between dosing and fluid sampling to decrease variability among subjects and timepoints. The active anabolite of 3TC, 3TC 5’-triphosphate (3TC-TP), has a relatively longer half-life of approximately 15 hours within cells^30^. While analysis of 3TC-TP would provide a quantitative assessment of active drug in each participant, this approach requires measurements of drug within isolated cells, which is a challenge for quantification in CSF.

While our study was not powered to assess efficacy, significant changes were detected in biomarkers related to neurodegeneration and neuroinflammation. In longitudinal studies, low levels of plasma Aβ42/40 and high levels of plasma pTau181, GFAP, and NfL are associated with subsequent cognitive decline^37^. In the current analysis, we find significant changes in fluid levels of Aβ42/40 and GFAP, and trending changes in NfL after six months of 3TC treatment. Plasma Aβ42/40 is inversely correlated with amyloid burden in the brain such that individuals with positive amyloid PET have significantly lower levels of Aβ42/40^40,41^. A lower Aβ42/40 ratio is also associated with amnestic mild cognitive impairment, and individuals with lower baseline levels of Aβ42/40 have increased risk of progression to dementia^42,43^. We find significant elevation of Aβ42/40 in plasma after six months of 3TC treatment compared to baseline, suggestive of a potentially protective effect of 3TC. In line with these data, we detect significant reduction of GFAP in CSF after six months of 3TC treatment. GFAP is a marker of reactive astrogliosis that is elevated in plasma and CSF of patients with preclinical, prodromal, and Alzheimer’s disease dementia^37,42,44–46^. The trending decrease in CSF levels of NfL, a marker of axonal damage that is elevated in CSF^47^ and plasma^48^ of individuals with mild cognitive impairment, Alzheimer’s dementia, and other neurodegenerative disorders^49^, further suggests a benefit due to 3TC treatment. Among neuroinflammatory biomarkers, we detect reduction of CSF Flt-1, also known as vascular endothelial growth factor receptor 1, and significant elevation of the proinflammatory cytokine IL-15 in plasma. Longitudinal biomarker analyses indicate that Flt-1 and IL-15 are elevated within CSF at preclinical, prodromal and dementia stages of Alzheimer’s disease, and are associated with cortical thinning and subsequent cognitive decline^38^. While analyses of plasma levels of IL-15 in individuals with Alzheimer’s disease are limited, a smaller study of 20 subjects with Alzheimer’s disease compared to 15 controls reports that IL-15 is slightly but significantly reduced in plasma of individuals with Alzheimer’s disease^50^; another study of 52 subjects with Alzheimer’s disease compared to 18 controls found no difference in IL-15 in CSF or plasma^51^. Taken together, our fluid biomarker analysis points toward compelling potential benefits of 3TC in individuals with mild cognitive impairment due to suspected Alzheimer’s disease.

Based on the similarities between exogenous retroviruses and retrotransposons, numerous studies have investigated the therapeutic efficacy of antiviral medications, including nucleoside analog reverse transcriptase inhibitors, to suppress retrotransposon activation. The Lighthouse trial, for example, which tested the effects of Triumeq in participants with ALS, has completed phase 2a^52^. Triumeq consists of two reverse transcriptase inhibitors, abacavir and 3TC, and an integrase inhibitor, dolutegravir, which blocks episomal DNA integration into genomic DNA. 35 out of 40 subjects completed the 24-week study. Non-serious adverse events included nausea and rash. While five deaths were expected based on modeling, only one death was observed five months after conclusion of the trial. The ALS functional rating scale demonstrated a declining trend. The trial is currently recruiting for phase 3, in which overall survival and disease progression will be measured in a larger cohort. An additional trial (NCT02437110) for a retroviral combination therapy (darunavir, ritonavir, dolutegravir, and tenofovir alafenamide) in ALS is currently in phase 1. A 12-month trial (NCT02363452) in children with Aicardi-Goutières Syndrome, in which the body mounts a viral response due to activation of retrotransposons, reports that participants administered 3TC, zidovudine, and abacavir have a significantly reduced blood interferon score after treatment^53^. Recent study results from a phase 2 trial in subjects with progressive supranuclear palsy led by Transposon Therapeutics report reduction in NfL, IL-6, and osteopontin after treatment with the reverse transcriptase inhibitor TPN-101, as well as stabilization of symptoms (NCT04993768). The phase 1 study LINE-AD, in which the nucleoside analog reverse transcriptase inhibitor emtricitabine is being tested in participants with mild cognitive impairment or Alzheimer’s disease (NCT04500847), is ongoing.

We find that reverse transcriptase activity in plasma is highly variable among participants and is unchanged to due 3TC treatment. Analysis of circulating levels of reverse transcriptase has several caveats. First, as the EnzCheck assay was performed on cell-free plasma, it is likely that levels of secreted reverse transcriptase do not reflect the level of reverse transcriptase activity within cells in circulation or within cells of the brain. Second, any type of double-stranded nucleic acid, whether it is produced via reverse transcription or another mechanism, will be detected by the assay. The Lighthouse trial (NCT02868580), an open label, multi-center study to investigate the safety and tolerability of Triumeq (3TC, abacavir and dolutegravir) in subjects with amyotrophic lateral sclerosis (ALS), analyzed DNA copy number of human endogenous retrovirus K (HERV-K) elements as a measure of target engagement^52^. While analysis of retrotransposon DNA copy number could potentially be leveraged in future studies of 3TC in Alzheimer’s disease and related tauopathies, the normalization of retrotransposon DNA copies, which are highly abundant and repetitive within the human genome, to a single-copy control gene using quantitative or digital PCR-based methods is a technical caveat of this approach.

In summary, we present findings from the first clinical trial testing the effects of reverse transcriptase inhibition in participants with mild cognitive impairment due to suspected Alzheimer’s disease. We find that 3TC is safe and well-tolerated among participants, and gains access to the CNS. We further detect beneficial effects of 3TC treatment on biomarkers associated with neurodegeneration and neuroinflammation, strongly supporting conduct of a larger, placebo-controlled study of 3TC in older adults with Alzheimer’s disease and, potentially, primary tauopathies.

## Data availability

The data required for interpretation, verification, and extension of the findings in the article are provided in the manuscript and Extended Data.

## Data Availability

All data produced in the present work are contained in the manuscript

## Acknowledgements

This study was funded by the Owens Foundation (BF) with support from the San Antonio Claude D. Pepper Older Americans Independence Center (P30 AG044271) (MLS, SG, SE, NM). Philanthropic support was provided by the Zachry Foundation. BF was supported by RF1 NS112391. ACS, EZ, GE, CW, RG and SS were supported by the South Texas Alzheimer’s Disease Research Center (P30 AG066546). GZ was supported by T32 GM113896. We thank Dr. Mourad Tighiouart and Dr. Nick Tatonetti for statistical advice.

## Author Contributions

BF, ACS, NM and GZ conceived the study. DAG and ACS developed the cognitive battery; ACS performed cognitive assessments. Statistical analyses were performed by PR, RF, CW, ACS and BF. JL and EG performed fluid sample processing. Biomarker and pharmacological analyses were performed by KP and MLC, respectively. SG was the study coordinator. SG, EG, AP, LD and SE participated in participant recruitment, trial coordination and execution. EZ, GE and SS performed lumbar punctures. BF prepared figures. The manuscript was written by ACS, GZ, KP, MLC, GE, NM and BF. All authors approved the final manuscript.

## Competing interests

BF serves on the Scientific Advisory Board for Transposon Therapeutics. All other authors have no competing interests to declare.

---

**Extended Data Figure 1.**
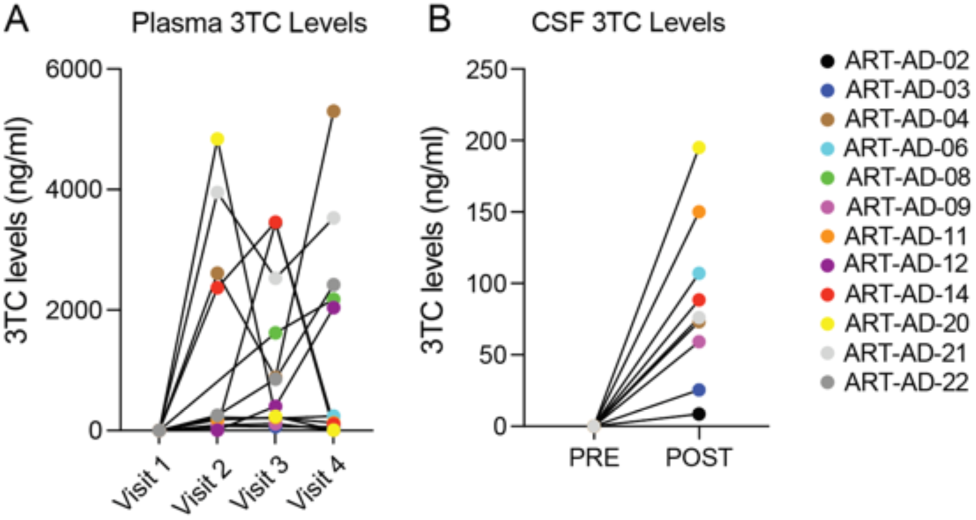
Levels of 3TC in plasma and CSF based on HPLC/MS/MS. Plasma, N=12; CSF, N=9.

**Extended Data Figure 2.**
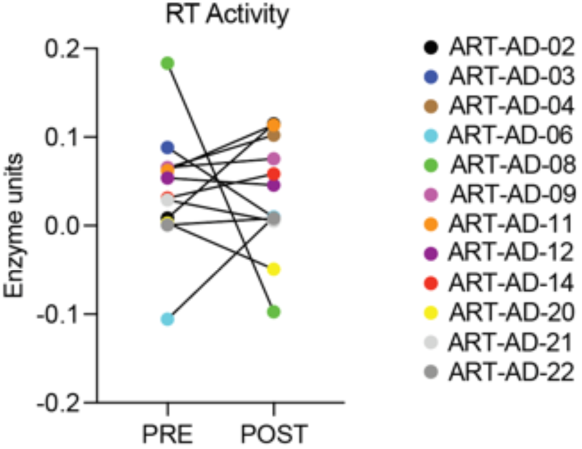
Reverse transcriptase activity in plasma. Reverse transcriptase activity was quantified using the EnzCheck Reverse Transcriptase Activity assay. N=12.

**Extended Data Figure 3.**
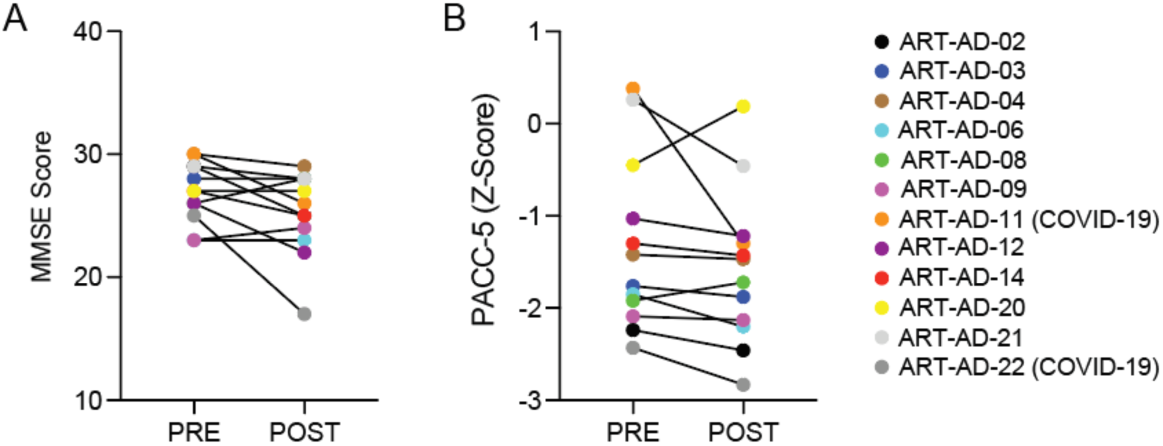
Cognitive scores per participant. A) MMSE score and B) PACC-5 Z-Scores for each participant at baseline and after 24 weeks of 3TC treatment. N=12.

**Extended Data Figure 4.**
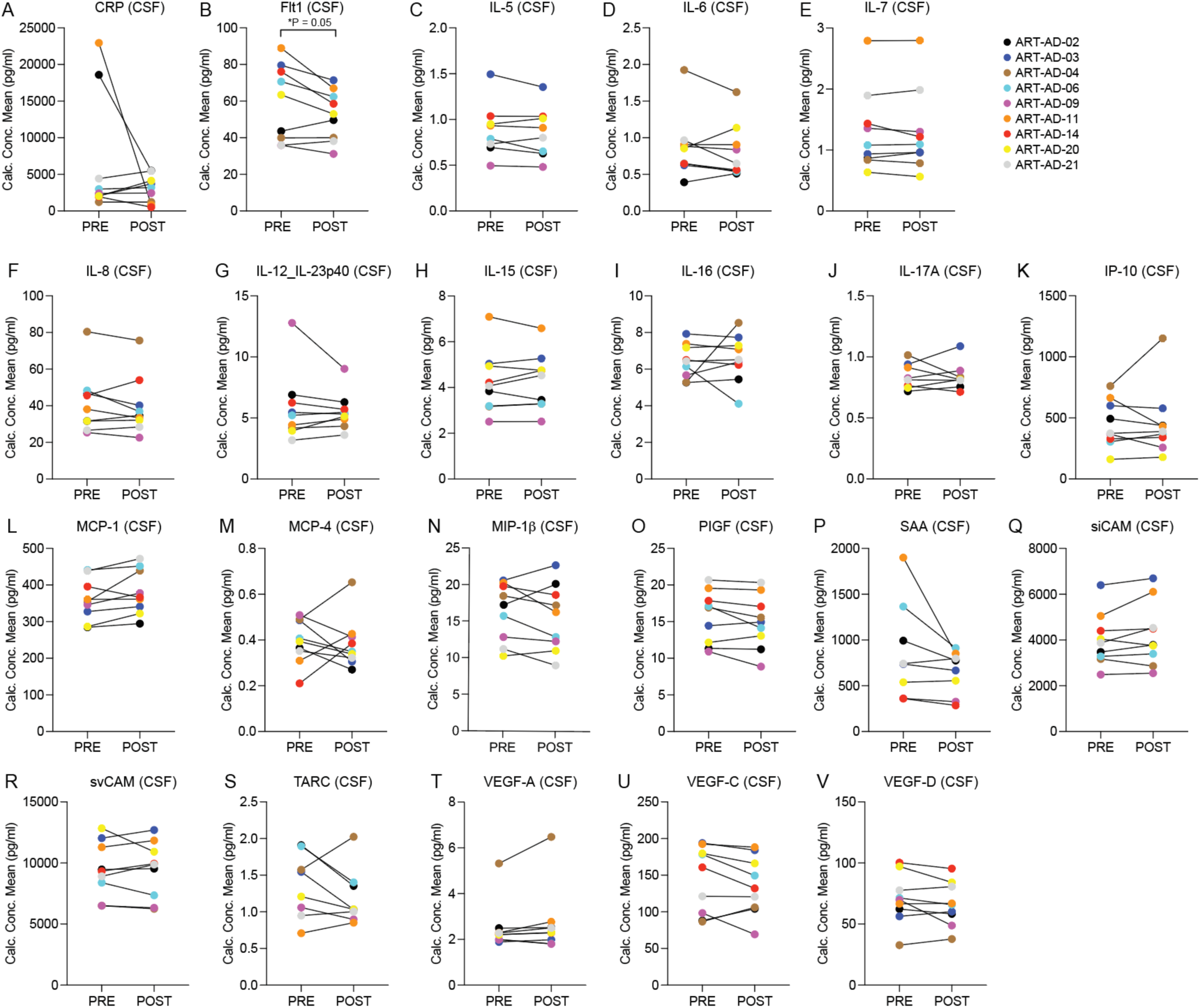
Neuroinflammatory biomarkers in CSF. The MSD platform was used to detect the indicated proteins in CSF of participants at baseline (PRE) and after 24 weeks of 3TC treatment (POST). Normality testing was based on the POST-PRE difference for each target. N=9. Significant *P* value is noted; all other *P* values are provided in Table 4. *P* values are based on two-sided paired sample t-test (normal distribution) or Wilcoxon matched-pairs signed rank test (non-normal distribution).

**Extended Data Figure 5.**
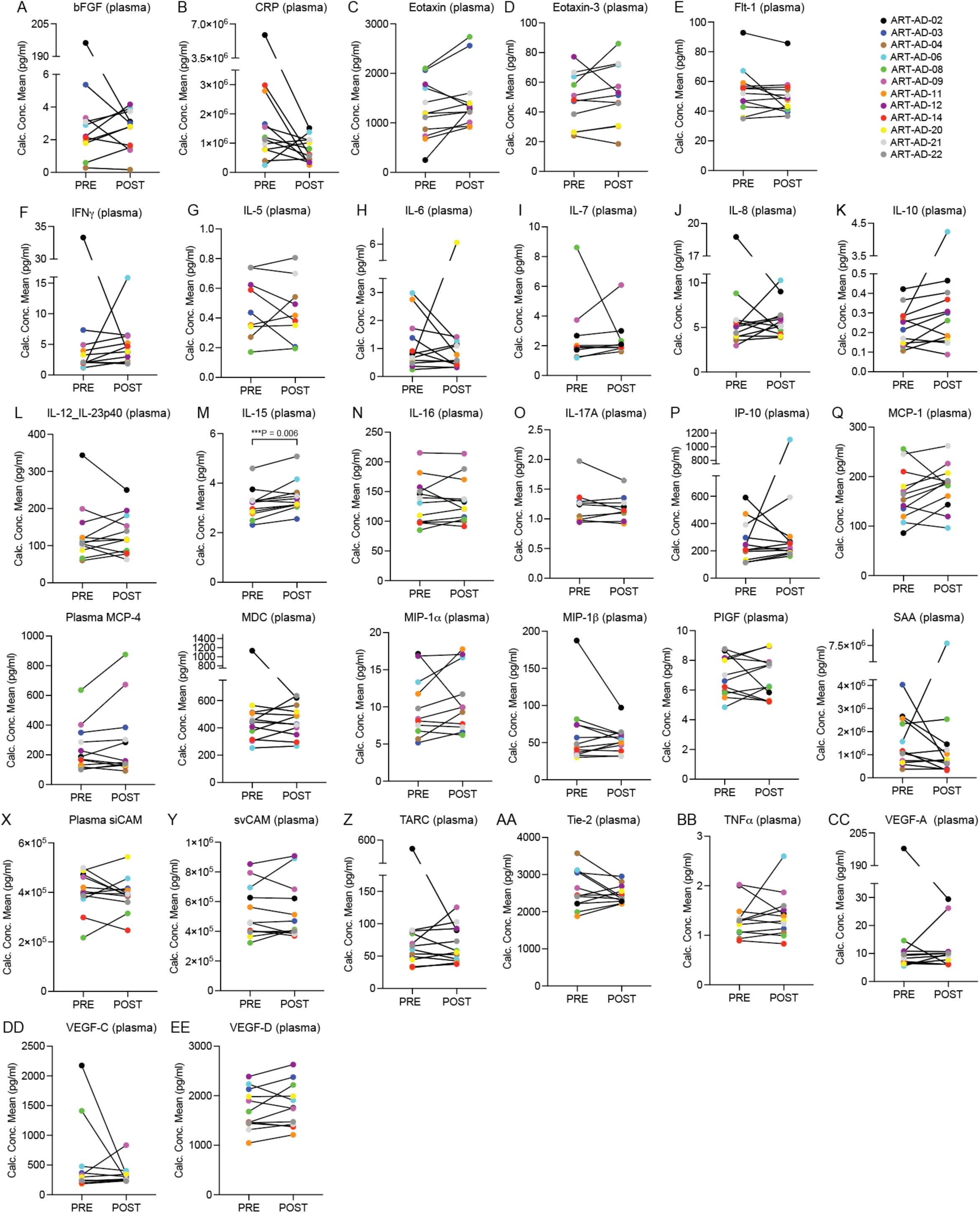
Neuroinflammatory biomarkers in plasma. The MSD platform was used to detect the indicated proteins in plasma of participants at baseline (PRE) and after 24 weeks of 3TC treatment (POST). Normality testing was based on the POST-PRE difference for each target. N=12. Significant *P* value is noted; all other *P* values are provided in Table 4. *P* values are based on two-sided paired sample t-test (normal distribution) or Wilcoxon matched-pairs signed rank test (non-normal distribution).

**Extended Data Table 1.**
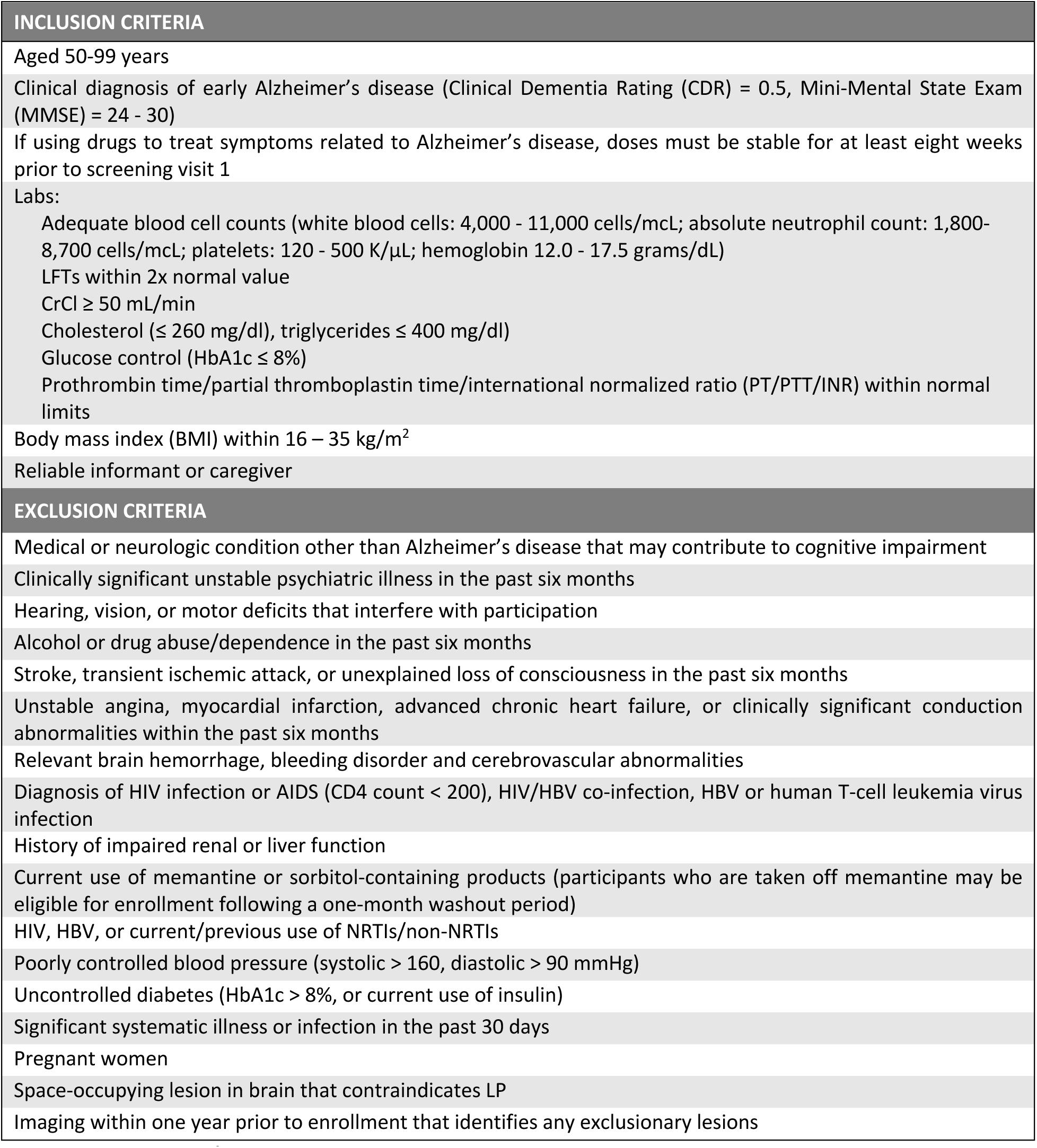
Eligibility criteria.

**Extended Data Table 2.**
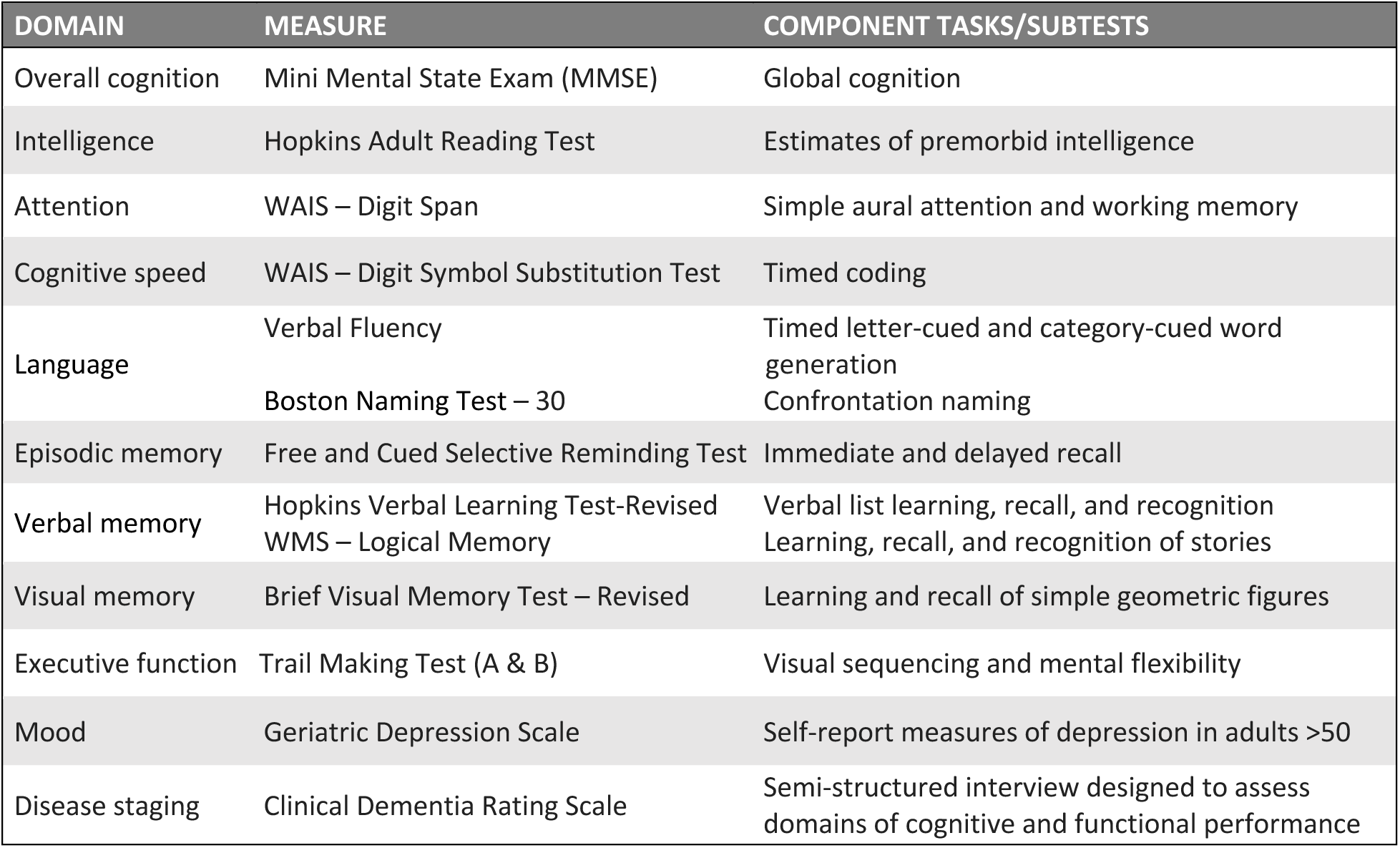
Neuropsychological battery.

## Notes

### Competing Interest Statement

BF serves on the Scientific Advisory Board of Transposon Therapeutics.

### Clinical Trial

NCT04552795

### Author Declarations

The Institutional Review Board of the University of Texas Health San Antonio gave ethical approval for this work.

## Citations

1. Wimo, A. et al. The worldwide costs of dementia 2015 and comparisons with 2010. Alzheimers Dement 13, 1–7 (2017).

2. Gustavsson, A. et al. Global estimates on the number of persons across the Alzheimer’s disease continuum. Alzheimers Dement 19, 658–670 (2023).

3. Haass, C. & Selkoe, D. If amyloid drives Alzheimer disease, why have anti-amyloid therapies not yet slowed cognitive decline? PLoS Biol 20, e3001694 (2022).

4. Shulman, M. et al. TANGO: a placebo-controlled randomized phase 2 study of efficacy and safety of the anti-tau monoclonal antibody gosuranemab in early Alzheimer’s disease. doi:10.1038/s43587-023-00523-w.

5. Monteiro, C. et al. Randomized Phase II Study of the Safety and Efficacy of Semorinemab in Participants With Mild-to-Moderate Alzheimer Disease: Lauriet. Neurology 101, E1391–E1401 (2023).

6. Braak, H. & Braak, E. Frequency of Stages of Alzheimer-Related Lesions in Different Age Categories. Neurobiol Aging 18, 351–357 (1997).

7. Sun, W., Samimi, H., Gamez, M., Zare, H. & Frost, B. Pathogenic tau-induced piRNA depletion promotes neuronal death through transposable element dysregulation in neurodegenerative tauopathies. Nat Neurosci 21, 1038–1048 (2018).

8. Ramirez, P. et al. Pathogenic tau accelerates aging-associated activation of transposable elements in the mouse central nervous system. Prog Neurobiol 208, 102181 (2022).

9. Ochoa, E. et al. Pathogenic tau-induced transposable element-derived dsRNA drives neuroinflammation. Sci Adv 9, eabq5423 (2023).

10. Guo, C. et al. Tau Activates Transposable Elements in Alzheimer’s Disease. CellReports 23, 2874– 2880 (2018).

11. Vallés-Saiz, L., Ávila, J. & Hernández, F. Lamivudine (3TC), a Nucleoside Reverse Transcriptase Inhibitor, Prevents the Neuropathological Alterations Present in Mutant Tau Transgenic Mice. Int J Mol Sci 24, (2023).

12. Wahl, D. et al. The reverse transcriptase inhibitor 3TC protects against age-related cognitive dysfunction. Aging Cell e13798 (2023) doi:10.1111/acel.13798.

13. Tam, O. H., Ostrow, L. W. & Gale Hammell, M. Diseases of the nERVous system: Retrotransposon activity in neurodegenerative disease. Mob DNA 10, 32 (2019).

14. Jönsson, M. E. et al. Activation of endogenous retroviruses during brain development causes an inflammatory response. EMBO J 40, (2021).

15. Chuong, E. B., Elde, N. C. & Feschotte, C. Regulatory activities of transposable elements: From conflicts to benefits. Nature Reviews Genetics Preprint at 10.1038/nrg.2016.139 (2017).

16. De Cecco, M. et al. L1 drives IFN in senescent cells and promotes age-associated inflammation. Nature 566, 73–78 (2019).

17. Gamdzyk, M. et al. cGAS/STING Pathway Activation Contributes to Delayed Neurodegeneration in Neonatal Hypoxia-Ischemia Rat Model: Possible Involvement of LINE-1. Mol Neurobiol (2020) doi:10.1007/s12035-020-01904-7.

18. Simon, M. et al. LINE1 Derepression in Aged Wild-Type and SIRT6-Deficient Mice Drives Inflammation. Cell Metab 29, 871–885.e5 (2019).

19. Zhao, K. et al. LINE1 contributes to autoimmunity through both RIG-I- and MDA5-mediated RNA sensing pathways. J Autoimmun 90, 105–115 (2018).

20. Gasior, S. L., Wakeman, T. P., Xu, B. & Deininger, P. L. The Human LINE-1 Retrotransposon Creates DNA Double-strand Breaks. J Mol Biol 357, 1383 (2006).

21. Macciardi, F. et al. A retrotransposon storm marks clinical phenoconversion to late-onset Alzheimer’s disease. Geroscience 44, 1525–1550 (2022).

22. Guo, C. et al. Tau Activates Transposable Elements in Alzheimer’s Disease. Cell Rep 5, 2874–2880 (2018).

23. Ochoa, E. et al. Pathogenic tau-induced transposable element-derived dsRNA drives neuroinflammation. Sci Adv 9, eabq5423 (2023).

24. Sun, Z. et al. Endogenous recapitulation of Alzheimer’s disease neuropathology through human 3D direct neuronal reprogramming. bioRxiv 2023.05.24.542155 (2023) doi:10.1101/2023.05.24.542155.

25. Cihlar, T., Birkus, G., Greenwalt, D. E. & Hitchcock, M. J. M. Tenofovir exhibits low cytotoxicity in various human cell types: Comparison with other nucleoside reverse transcriptase inhibitors. Antiviral Res 54, 37–45 (2002).

26. Ogedegbe, A. O. & Sulkowski, M. S. Antiretroviral-associated liver injury. Clin Liver Dis 7, 475–499 (2003).

27. van Leeuwen, R. et al. The safety and pharmacokinetics of a reverse transcriptase inhibitor, 3TC, in patients with HIV infection: a phase I study. AIDS 6, 1471–1476 (1992).

28. Quercia, R. et al. Twenty-five years of lamivudine: Current and future use for the treatment of HIV-1 infection. J Acquir Immune Defic Syndr (1988) 78, 125–135 (2018).

29. Magagnoli, J. et al. Reduction of human Alzheimer’s disease risk and reversal of mouse model cognitive deficit with nucleoside analog use. doi:10.1101/2023.03.17.23287375.

30. Else, L. J. et al. Pharmacokinetics of Lamivudine and Lamivudine-Triphosphate after Administration of 300 Milligrams and 150 Milligrams Once Daily to Healthy Volunteers: Results of the ENCORE 2 Study. (2012) doi:10.1128/AAC.05599-11.

31. Chang, A., Ostrove, J. M. & Bird, R. E. Development of an improved product enhanced reverse transcriptase assay. J Virol Methods 65, 45–54 (1997).

32. Lebensztejn, D. M. & Kaczmarski, M. Lamivudine-associated thrombocytopenia. Am J Gastroenterol 97, 2687–2688 (2002).

33. Ormseth, E. J. et al. Hepatic Decompensation Associated With Lamivudine: A Case Report and Review of Lamivudine-Induced Hepatotoxicity. American Journal of Gastroenterology 96, 1619– 1622 (2001).

34. Kakubu, M. A. M., Bikinesi, T. & Katoto, P. D. M. C. Lamivudine induced pure red cell aplasia and HIV-1 drug resistance-associated mutations: a case report. Oxf Med Case Reports 2023, 106–109 (2023).

35. Bosch, B. et al. Weight and Metabolic Changes After Switching From Tenofovir Alafenamide/Emtricitabine (FTC) +Dolutegravir (DTG), Tenofovir Disoproxil Fumarate (TDF)/FTC + DTG, and TDF/FTC/Efavirenz to TDF/Lamivudine/DTG. Clinical Infectious Diseases ® 76, 1492– 1497 (2022).

36. Papp, K. V., Rentz, D. M., Orlovsky, I., Sperling, R. A. & Mormino, E. C. Optimizing the preclinical Alzheimer’s cognitive composite with semantic processing: The PACC5. Alzheimer’s and Dementia: Translational Research and Clinical Interventions (2017) doi:10.1016/j.trci.2017.10.004.

37. Chatterjee, P. et al. Plasma Aβ42/40 ratio, p-tau181, GFAP, and NfL across the Alzheimer’s disease continuum: A cross-sectional and longitudinal study in the AIBL cohort. Alzheimer’s and Dementia 19, 1117–1134 (2023).

38. Janelidze, S. et al. CSF biomarkers of neuroinflammation and cerebrovascular dysfunction in early Alzheimer disease. Neurology 91, e867–e877 (2018).

39. Johnson, M. A., Moore, K. H. P., Yuen, G. J., Bye, A. & Pakes, G. E. Clinical pharmacokinetics of lamivudine. Clinical Pharmacokinetics Preprint at 10.2165/00003088-199936010-00004 (1999).

40. Schindler, S. E. et al. High-precision plasma β-amyloid 42/40 predicts current and future brain amyloidosis. (2019) doi:10.1212/WNL.0000000000008081.

41. Rembach, A. et al. Changes in plasma amyloid beta in a longitudinal study of aging and Alzheimer’s disease. Alzheimer’s and Dementia 10, 53–61 (2014).

42. Verberk, I. M. W. et al. Plasma Amyloid as Prescreener for the Earliest Alzheimer Pathological Changes. Ann Neurol 84, 648–658 (2018).

43. Pérez-Grijalba, V. et al. Plasma Aβ42/40 Ratio Detects Early Stages of Alzheimer’s Disease and Correlates with CSF and Neuroimaging Biomarkers in the AB255 Study. J Prev Alzheimers Dis 6, 34–41 (2019).

44. Fukuyama, R., Izumoto, T. & Fushiki, S. The Cerebrospinal Fluid Level of Glial Fibrillary Acidic Protein Is Increased in Cerebrospinal Fluid from Alzheimer’s Disease Patients and Correlates with Severity of Dementia. Eur Neurol 46, 35–38 (2001).

45. Jesse, S. et al. Glial Fibrillary Acidic Protein and Protein S-100B: Different Concentration Pattern of Glial Proteins in Cerebrospinal Fluid of Patients with Alzheimer’s Disease and Creutzfeldt-Jakob Disease. Journal of Alzheimer’s Disease 17, 541–551 (2009).

46. Oeckl, P. et al. Glial Fibrillary Acidic Protein in Serum is Increased in Alzheimer’s Disease and Correlates with Cognitive Impairment. Journal of Alzheimer’s Disease 67, 481–488 (2019).

47. Zetterberg, H. et al. Association of Cerebrospinal Fluid Neurofilament Light Concentration With Alzheimer Disease Progression. JAMA Neurol 73, 60–67 (2016).

48. Mattsson, N. et al. Association of Plasma Neurofilament Light With Neurodegeneration in Patients With Alzheimer Disease. JAMA Neurol 74, 557 (2017).

49. Simrén, J., Ashton, N. J., Blennow, K. & Zetterberg, H. An update on fluid biomarkers for neurodegenerative diseases: recent success and challenges ahead. Current Opinion in Neurobiology Preprint at 10.1016/j.conb.2019.11.019 (2020).

50. Rentzos, M. et al. Circulating interleukin-15 in dementia disorders. Journal of Neuropsychiatry and Clinical Neurosciences 19, 318–325 (2007).

51. Johansson, P. et al. Reduced cerebrospinal fluid concentration of interleukin-12/23 subunit p40 in patients with cognitive impairment. (2017) doi:10.1371/journal.pone.0176760.

52. Gold, J. et al. Safety and tolerability of Triumeq in amyotrophic lateral sclerosis: the Lighthouse trial. Amyotroph Lateral Scler Frontotemporal Degener (2019) doi:10.1080/21678421.2019.1632899.

53. Rice, G. I. et al. Reverse-transcriptase inhibitors in the Aicardi-Goutières syndrome. New England Journal of Medicine Preprint at 10.1056/NEJMc1810983 (2018).

